# The prevalence and risk factors for phantom limb pain in people with amputations: a systematic review and meta-analysis

**DOI:** 10.1101/19008961

**Authors:** K. Limakatso, G. J. Bedwell, V. J. Madden, R. Parker

**Affiliations:** Pain Management Unit, Department of Anaesthesia and Perioperative Medicine, University of Cape Town, Cape Town, South Africa; Division of Physiotherapy, Department of Health and Rehabilitation Sciences, University of Cape Town, Cape Town, South Africa; Neuroscience Institute. University of Cape Town, Groote Schuur Hospital, Cape Town, South Africa

## Abstract

**Background and objective:** Phantom limb pain (PLP) is a common complication in people with limb amputations. There are conflicting reports in the literature regarding the prevalence of PLP in people with limb amputations. Therefore, this review aimed to determine the estimated pooled prevalence of PLP, and risk factors associated with this complication.

**Methods:** Articles published between 1980 and July 2019 were identified through a systematic search of the following electronic databases: MEDLINE/PubMed, PsycINFO, PsycArticles, Cumulative Index to Nursing and Allied Health Literature, Africa−Wide Information, Health Source: Nursing/Academic Edition, SCOPUS, Web of Science and Academic Search Premier. Grey literature was searched on databases for preprints. Two reviewers independently performed the screening of articles, data extraction and risk of bias assessment. The meta−analyses were conducted using the random−effects model. A statistically significant level for the analyses was set at p>0.05.

**Results:** The pooling of all studies demonstrated a prevalence estimate of 63% [95% CI: 58.23−67.05] with high heterogeneity [I2=95.70% (95% CI: 95.10−96.20)]. The prevalence of PLP was significantly lower in developing countries compared to developed countries [53.98% vs 64.55%; p=0.04]. Persistent pre−operative pain, proximal site of amputation, lower limb amputation, stump pain and phantom sensations were identified as risk factors for PLP.

**Conclusion:** This systematic review and meta−analysis estimates that six of every 10 people with an amputation report PLP − a high and important prevalence of PLP. Health care professionals ought to be aware of the high rates of PLP and implement strategies to reduce PLP by addressing known risk factors, specifically those identified by the current study.

## Background

Phantom limb pain (PLP) is commonly defined as pain felt in the missing portion of an amputated limb (Ehde, Czerniecki et al. 2000). Some cases of PLP have been reported in people with congenital amputations (Garcia, Flores et al. 2018). However, PLP commonly occurs in people with limb amputations due to trauma or surgery (Neil 2015). It has been proposed that risk factors such as persisting pre-operative pain, diabetic cause of amputation and prosthesis use contribute to the onset of PLP (Dijkstra, Geertzen et al. 2002). Phantom limb pain is common and often associated with negative consequences such as depression and problems with prosthesis use, sleep and participation in activities of daily function (Morgan, Friedly et al. 2017).

The prevalence of PLP among people with amputations is unclear, perhaps because of conflicting reports across the literature. While one study reported a high prevalence of 85.6% (Clark, Bowling et al. 2013), another reported a markedly lower prevalence of 0% (Kooijman, Dijkstra et al. 2000). The conflicting reports in the literature regarding the prevalence of PLP are perplexing, but may be due to variations in the time period during which the studies were undertaken, the countries in which the studies were conducted, and the recruitment processes used during collection of epidemiological data (Maimela, Alberts et al. 2016).

Low prevalence rates were reported by early studies conducted during a period when PLP was commonly characterised as a psychiatric disorder (Jensen, Krebs et al. 1985). The low prevalence rates could be explained by patients’ fear of reporting their pain to avoid the stigma associated with PLP (Weeks, Anderson-Barnes et al. 2010). Low prevalence rates were also recorded in studies conducted in developing countries where stigma associated with chronic pain conditions is common, and effective strategies to follow up patients after amputations are still lacking (Mishra, Bhatnagar et al. 2008, Gosselin, Gyamfi et al. 2011). Furthermore, many prevalence studies of PLP were conducted in clinical settings using patients continuing with medical care, introducing a selection bias (Sherman, Sherman et al. 1980, Desmond and MacLachlan 2010, Ventham, Heyburn et al. 2010, Byrne 2011, Ahmed, Bhatnagar et al. 2017). Therefore, patients without continuing clinical care may be unaccounted for in these statistics.

Prevalence studies are key to informing researchers, clinicians, policy-makers and the public about the burden of diseases in society (Moloi, Watkins et al. 2016). However, the burden of PLP is unclear because of a wide variation in the reported prevalence estimates. To our knowledge, no systematic review has synthesised data on the prevalence and risk factors for PLP. To address this gap in the literature, we conducted a systematic review and meta-analysis by gathering and critically appraising relevant literature regarding the prevalence and associated risk factors for PLP in people with amputations.

### Objective

The primary aim of this systematic review was to estimate the prevalence of PLP in people with amputations. The secondary aim was to determine whether there is a difference in the prevalence of PLP in developed and developing countries as per the World Economic Situation and Prospects classification system (WESP 2018). The exploratory aim was to identify associated risk factors for PLP in people with limb amputations.

## MATERIAL AND METHODS

This systematic review was developed in accordance with the Preferred Reporting Items of Systematic Reviews and Meta-Analysis (PRISMA) guidelines (Moher, Shamseer et al. 2015). The review protocol was registered on PROSPERO - an international prospective register of systematic reviews with health-related outcome, and published in Systematic Reviews (Limakatso, Bedwell et al. 2019). The PRISMA criteria fulfilled by this systematic review are presented in additional file 1.

### Data sources and search procedure

The lead investigator (KL) and a senior librarian (MS) developed a comprehensive search strategy (Appendix A) using five Medical Subject Headings (MeSH): prevalence, risk factors, amputation, phantom limb and epidemiology. Articles published between 1980 and July 2019 were identified through a systematic search of the following electronic databases: MEDLINE/PubMed (via EBSCOhost), PsycINFO (via EBSCOhost), PsycArticles, Cumulative Index to Nursing and Allied Health Literature (CINAHL) (via EBSCOhost), Africa-Wide Information (via EBSCOhost), Health Source: Nursing/Academic Edition (via EBSCOhost), SCOPUS, Web of Science and Academic Search Premier (via EBSCOhost). Grey literature was searched on bioRxiv (www.biorxiv.org), Preprints (www.preprints.org), Open Science Framework (www.osf.io) and medRxiv (www.medrxiv.org). The reference lists of eligible studies were searched manually to identify more studies that may have been eligible for inclusion in this review. Studies identified from the literature search were saved using the citation manager software programme (EndNote x8), which was also used to remove duplicates (Rathvon 2017).

### Study selection

We included case-control, cross-sectional and cohort studies that investigated the prevalence of PLP in surgical, traumatic and congenital upper or lower limb amputees who were 18 years or older. The risk factors for PLP were identified from the included studies. Only studies written in English, with full text available, were eligible for inclusion in this review. We excluded studies that did not meet the inclusion criteria. Two reviewers (KL and GJB) independently screened study titles and abstracts for eligibility. Studies identified in the initial screening as potentially eligible were assessed for eligibility in full-text form by the same reviewers, using the inclusion/exclusion criteria. The study selection procedure was performed using a Microsoft Excel spreadsheet (2016) on which the studies were listed and marked as either eligible or ineligible. A PRISMA flow diagram (Fig 1) represents the entire screening process detailing the numbers of included and excluded studies, with reasons for exclusion. After each stage, results were compared, and disagreements resolved through discussion.

**Fig 1.**
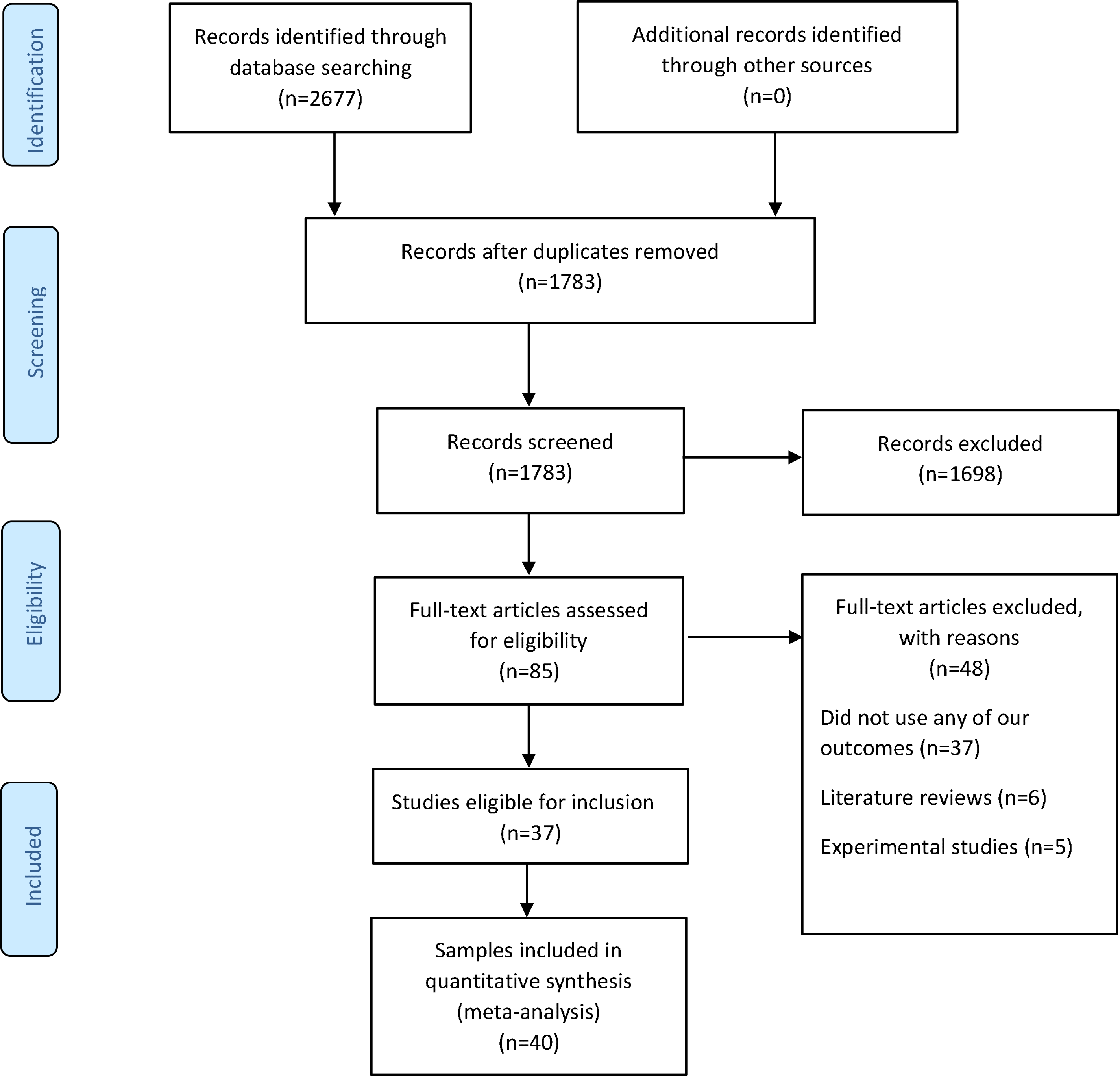
The PRISMA flow diagram illustrating the screening process.

### Risk of bias assessment

Two reviewers (KL and GJB) independently assessed the included articles for risk of bias using a risk of bias assessment tool for prevalence studies that had been developed by Hoy et al (Hoy, Brooks et al. 2012). This tool assesses the risk of bias based on 10 categories which evaluate the study’s external and internal validity. Each category of the risk of bias tool was set as “high risk” if the study scored “high risk” for any single item within that category, and “low risk” if it scored “low risk” for all items in that category. Categories with “low risk” and “high risk” were given a rating of zero and one respectively. The summary risk of bias rating for each study was presented as “low risk” (score: 0-3), “moderate risk” (score: 4-6), or “high risk” (score: 7-10).

### Data extraction

Two reviewers (KL and GJB) used a pre-piloted customised data extraction sheet to independently extract relevant data from included studies. Data extracted included: the names of authors, year of study publication, study design and setting, country of study, sample size, participants’ age and sex, site of amputation, method of data collection, PLP prevalence (%), as well as risk factors and their measures of association with PLP. The developmental status of the countries in which the studies were conducted was identified using the World Economic Situation and Prospects classification system (WESP 2018).

### Data analysis

Data extracted from individual studies were entered into an Excel spreadsheet for analysis. All meta-analyses were conducted using Open Meta Analyst software available on (http://www.cebm.brown.edu/openmeta). In this, we deviated from the registered protocol, which specified the use of Review Manager 5, because Review Manager 5 is not suitable for conducting meta-analyses of single arm studies. Cohen’s Kappa was used to report inter-rater agreement during screening, data extraction and risk of bias assessment, and can be interpreted as minimal (0-0.39), weak (0.40-0.59), moderate (0.60-0.79) or strong (0.80-0.90) (Cohen 1960). Clinical heterogeneity was evaluated qualitatively, based on similarities or differences in participant and outcome characteristics, recruitment and assessment procedures, and study setting (Gagnier, Moher et al. 2012). Statistical heterogeneity was assessed using the *I*^*2*^ statistic, and the results were presented as low (<25%), moderate (25-50%) and high (>50%) (Higgins, Thompson et al. 2003). Subject to consideration of heterogeneity and risk of bias, studies were pooled for meta-analysis using a random effects model to determine a sample-weighted summary estimate of PLP prevalence across all included studies. A funnel plot was generated to assess for possible publication bias. In addition, the Egger’s test was conducted to assess the funnel plot for asymmetry (Egger, Smith et al. 1997). To address high statistical heterogeneity, we deviated from protocol to sub-group studies into those with low risk of bias and those with moderate and high risk of bias, and conducted separate meta-analyses, and compared the two pooled estimates using a two-tailed Mann-Whitney U test. We also sub-grouped studies by the developmental status of the country in which each study had been conducted (WESP 2018), as planned in the protocol and, again, compared the estimates using a two-tailed Mann-Whitney U test. Potential risk factors for PLP were identified from the included studies and synthesised descriptively. When an association was confirmed, the strength of association between PLP and each risk factor was classified as either “weak”, “moderate”, “strong” or “very strong”, according to the guidelines for interpreting the strength of association in epidemiology studies (Rosenthal 1996, Nicolich and Gamble 2011, Healey 2014, Allen 2017). We calculated Phi (Ø) to determine the strength of association in studies that used the chi-square test as a measure of association. This sample-size-adjusted chi-square statistic has been shown to provide a more accurate reflection of the strength of association between two variables than the interpretation of chi-square and probability (P) values, where high chi-square and p values are thought to represent a strong association between variables (Frankfort-Nachmias and Leon-Guerrero 2017). Alpha was set at 0.05 for all analyses.

## Results

The initial literature search returned 2677 records, of which 1783 remained after the removal of duplicates. Initial screening of titles and abstracts identified 85 studies that were eligible for full-text review. Full-text review identified 37 studies that were eligible for inclusion in this review. Three of these studies reported two studies each (Kooijman, Dijkstra et al. 2000, Reiber, McFarland et al. 2010, Byrne 2011). Therefore, a total of 40 data sets were included in our analysis. The screening process reflected moderate agreement (Kappa=0.70) between reviewers.

### Study characteristics

The study characteristics are summarised in Table 1. The included studies had used cross-sectional (n=38) and cohort (n=2) study designs. Thirty-three of 40 studies had been conducted in developed countries (WESP 2018). Of these, the majority had been conducted in North America [USA (n=7); Canada (n=2)] and Europe [United Kingdom (n=8); Germany (n=5); Netherlands (n=4); Ireland (n=2); Poland (n=1)] (Fig 2). Only seven studies had been conducted in developing countries [Iran (n=2); Iraq (n=1); India (n=1); Brazil (n=1); Pakistan (n=1); Cambodia (n=1)]. Table 1 reflects the wide range of data collection approaches used by the studies. The included studies had been published between 1986 and 2019. The data extraction process had moderate agreement (Kappa=0.71) between reviewers prior to discussion.

**Table 1.** Summary of study and participant characteristics by study.

| Authors | Study type | Country of study | Development status | Method of data collection | Sample size | Age Mean (SD) | Sex M/F | Level of amputation (UL/LL) | PLP prevalence (%) |
| --- | --- | --- | --- | --- | --- | --- | --- | --- | --- |
| Ahmed et al., 2017 | Cross-sectional | India | Developing | Self-reported questionnaire | 139 | 38.23 (1.54) | 102/37 | 36/103 | 41 |
| Aldington et al., 2014 | Cross-sectional | UK | Developed | Self-reported questionnaire | 48 | 28.8 (6.7) | - | 11/54 | 49 |
| Bekrater et al., 2015 | Cross-sectional | Germany | Developed | Postal and telephone questionnaire | 3234 | 64.37 (15.89) | 2637/597 | 824/2410 | 62.55 |
| Bin Ayaz et al., 2015 | Cross-sectional | Pakistan | Developing | Face-to-face interview | 268 | 28 (6) | 266/2 | 35/233 | 42.5 |
| Bosmans et al., 2007 | Cross-sectional | Netherlands | Developed | Face-to-face interview | 16 | 66.5 (39-86)* | 11/5 | 0/16 | 81.25 |
| Buchanan et al., 1986 | Cross-sectional | Canada | Developed | Face-to-face interview | 716 | - | 616/100 | 43/647 | 62.4 |
| Byrne et al., 2011<sup>a</sup> | Cross-sectional | New Zealand | Developed | Face-to-face interview | 29 | 41.7 (4.8) | 25/4 | 7/24 | 69 |
| Byrne et al., 2011<sup>b</sup> | Cross-sectional | Cambodia | Developing | Face-to-face interview | 29 | 40.3 (10.5) | 25/4 | 1/28 | 51.7 |
| Clark et al., 2013 | Cross-sectional | UK | Developed | Postal and telephone questionnaire | 102 | 70.9 (1.27) | - | 0/97 | 85.6 |
| Datta et al., 2004 | Cohort | UK | Developed | Postal questionnaire | 60 | 58.1 (-) | 48/12 | 60/0 | 60 |
| Desmond et al., 2010 | Cross-sectional | Ireland | Developed | Self-reported questionnaire | 141 | 74.8 (-) | 138/3 | 141/0 | 42.6 |
| Dijkstra et al., 2002 | Cross-sectional | Netherlands | Developed | Postal questionnaire | 536 | - | 367/150 | 99/433 | 72 |
| Ehde et al., 2000 | Cross-sectional | USA | Developed | Postal questionnaire | 255 | 55.1 (14.3) | 207/48 | 0/255 | 72 |
| Ephraim et al., 2005 | Cross-sectional | USA | Developed | telephone interview | 914 | 50.3 (13.3) | 552/362 | 100/812 | 79.9 |
| Gallagher et al., 2001 | Cross-sectional | Ireland | Developed | Postal questionnaire | 104 | 45.3 (18.9) | 78/26 | 0/104 | 69.2 |
| Hanley et al., 2006 | Cross-sectional | USA | Developed | Postal and telephone questionnaire | 255 | 55 (14.3) | 207/48 | 0/255 | 72 |
| Hanley et al., 2009 | Cross-sectional | USA | Developed | Postal questionnaire | 104 | 46.9 (14.1) | 75/29 | 104/0 | 79 |
| Hnoosh et al., 2014 | Cross-sectional | Iraq | Developing | Self-reported questionnaire | 118 | 32 (12.9) | 97/21 | 0/181 | 61 |
| Houghton et al., 1994 | Cross-sectional | UK | Developed | Postal questionnaire | 176 | 71 (-) | - | 0/176 | 78 |
| Kern et al., 2012 | Cross-sectional | Germany | Developed | Postal questionnaire | 537 | 59 (-) | 382/155 | 24/513 | 74.5 |
| Ketz et al., 2008 | Cross-sectional | Germany | Developed | Self-reported questionnaire | 30 | - | 30/0 | 7/27 | 77 |
| Kooijman et al., 2000<sup>a</sup> | Cross-sectional | Netherlands | Developed | Unclear | 72 | 44.2 (35-65)* | 57/15 | 72/0 | 51 |
| Kooijman et al., 2000<sup>b</sup> | Cross-sectional | Netherlands | Developed | Unclear | 27 | 30.5 (9.3-53)* | 13/13 | 27/0 | 0 |
| Larbig et al., 2019 | Cohort | Germany | Developed | Face-to-face interview and self-reported questionnaire | 52 | - | 41/11 | 2/50 | 75 |
| Morgan et al., 2017 | Cross-sectional | USA | Developed | Self-reported and internet questionnaire | 1296 | 54.4 (13.7) | 909/387 | 0/1296 | 48.1 |
| Noguchi et al., 2019 | Cross-sectional | Japan | Developed | Medical records | 44 | - | 33/11 | 22/22 | 50 |
| Penna et al., 2018 | Cohort | Australia | Developed | Medical records | 96 | - | 74/22 | 0/96 | 52.2 |
| Probstner et al., 2010 | Cross-sectional | Brazil | Developing | Self-reported questionnaire | 75 | 54.4 (18.5) | 50/25 | 6/69 | 46.7 |
| Rafferty et al., 2015 | Cross-sectional | UK | Developed | Self-reported questionnaire | 75 | 26.3 (18-42)* | 74/1 | 0/84 | 85 |
| Rahimi et al., 2012 | Cross-sectional | Iran | Developing | Face-to-face interview | 335 | 42.1 (6.32) | 324/11 | 0/670 | 66.7 |
| Rayegani et al., 2010 | Cross-sectional | Iran | Developing | Face-to-face interview and self-reported questionnaire | 335 | - | 327/8 | 0/670 | 64 |
| Razmus et al., 2017 | Cross-sectional | Poland | Developed | Face-to-face interview and self-reported questionnaire | 22 | 61 (11.3) | 15/7 | 3/22 | 59 |
| Reiber et al., 2010<sup>a</sup> | Cross-sectional | USA | Developed | Postal, internet and telephone questionnaire | 298 | 60.7 (3.0) | 298/0 | 78/300 | 72.2 |
| Reiber et al., 2010<sup>b</sup> | Cross-sectional | USA | Developed | Postal, internet and telephone questionnaire | 283 | 29.3 (5.8) | 274/9 | 78/273 | 76 |
| Resnik et al., 2019 | Cross-sectional | Canada | Developed | Telephone interview | 808 | 63.2 (14.2) | 787/21 | 840/0 | 76.1 |
| Richardson et al., 2007 | Cohort | UK | Developed | Face-to-Face interview | 59 | 63.8 (10.4) | 37/22 | 0/59 | 78.8 |
| Richardson et al., 2015 | Cross-sectional | UK | Developed | Face-to-face interview | 89 | 65.5 (11.4) | 64/25 | 0/89 | 63 |
| Schley et al., 2008 | Cross-sectional | Germany | Developed | Postal and telephone questionnaire | 65 | 45 (18-80)* | 60/5 | 65/0 | 44.6 |
| Wartan et al., 1997 | Cross-sectional | UK | Developed | unclear | 526 | 73 (-)* | 526/0 | 99/471 | 62 |
| Yin et al., 2017 | Cross-sectional | China | Developed | Telephone interview | 391 | - | - | - | 29 |

\* Indicates the median age and range
- The number of amputations and males versus females do not add up to the total sample size because some participants had more than one amputation and these data were not provided.

**Fig 2.**
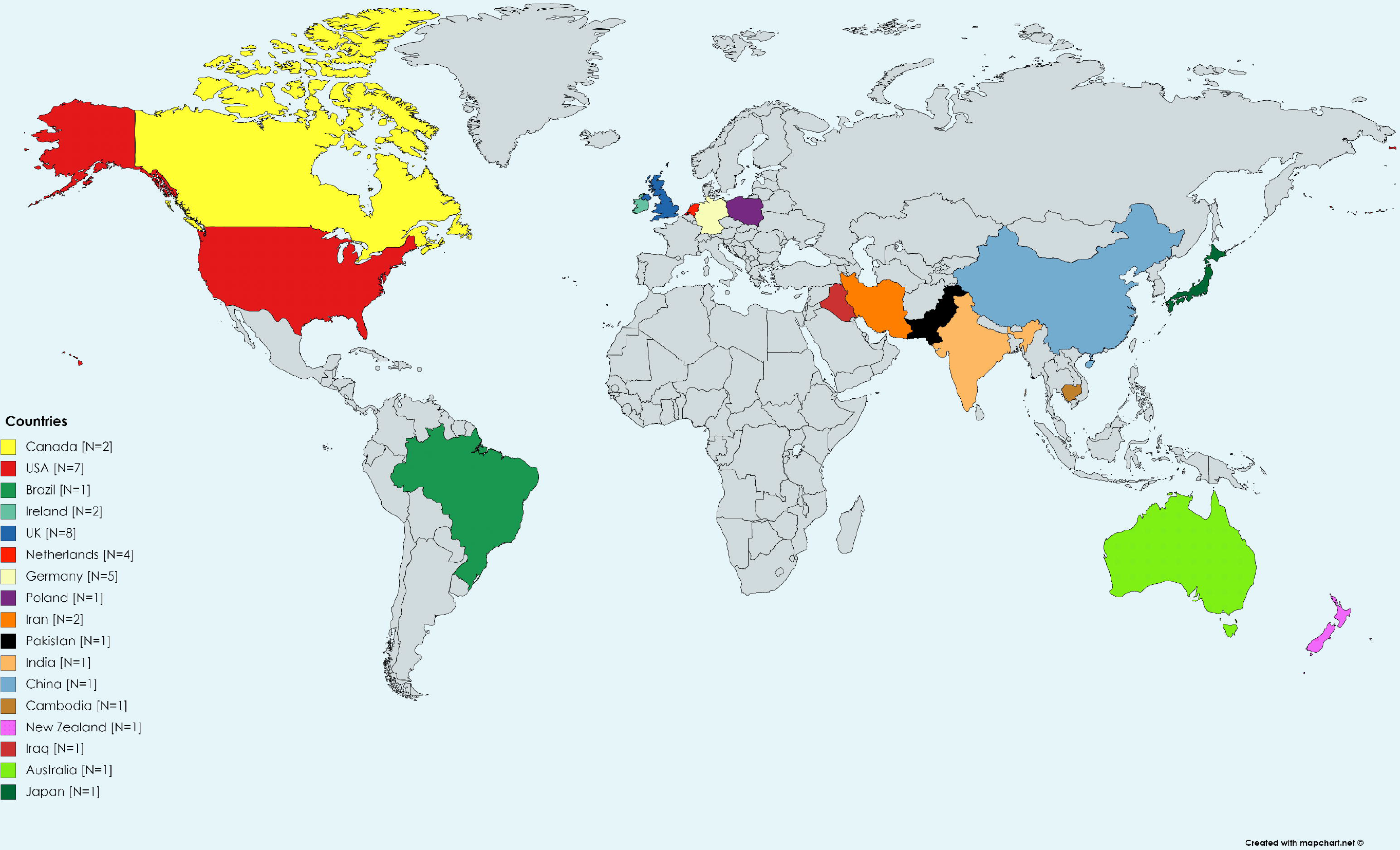
World map showing the countries in which the studies had been conducted.

### Participant characteristics

The eligible studies provided data from a total of 12765 participants (9826 male; 2196 female) who had undergone upper limb (n=2784) and lower limb (n=10539) amputations. Participant characteristics are provided in Table 1.

### Risk of bias assessment

The risk of bias assessment revealed moderate agreement (Kappa=0.69) between reviewers prior to discussion. The results of the risk of bias assessment are reported in Table 2. Four studies had an overall rating of “low risk” (Rayegani, Aryanmehr et al. 2010, Rahimi, Mousavi et al. 2012, Aldington, Small et al. 2014, Hnoosh 2014). Six studies scored “low risk” for selection bias, for using a sample that was a close representation of the national population (Ephraim, Wegener et al. 2005, Kern, Busch et al. 2009, Rahimi, Mousavi et al. 2012, Hnoosh 2014, Bekrater-Bodmann, Schredl et al. 2015, Morgan, Friedly et al. 2017). Eight studies scored “low risk” for study participation bias, because their response rates for participation were ≥75% (Wartan, Hamann et al. 1997, Kooijman, Dijkstra et al. 2000, Datta, Selvarajah et al. 2004, Richardson, Glenn et al. 2006, Rayegani, Aryanmehr et al. 2010, Rahimi, Mousavi et al. 2012, Rafferty, Bennett Britton et al. 2015, Morgan, Friedly et al. 2017). Twelve studies scored “low risk” for measurement bias, for using a clear definition of PLP (Ehde, Czerniecki et al. 2000, Dijkstra, Geertzen et al. 2002, Ephraim, Wegener et al. 2005, Hanley, Ehde et al. 2006, Ketz 2008, Desmond and MacLachlan 2010, Rayegani, Aryanmehr et al. 2010, Aldington, Small et al. 2014, Hnoosh 2014, Bekrater-Bodmann, Schredl et al. 2015, Bin Ayaz, Ikram et al. 2015, Noguchi, Saito et al. 2019). Other studies scored “high risk” for measurement bias, for not providing a clear definition of PLP (e.g pain felt in the limb after amputation). All the studies scored “high risk” for measurement bias, for using an instrument that has not been shown to be valid and reliable for measuring the outcome of interest. However, all studies scored “low risk” for reporting bias, for appropriately reporting the numerators and denominators for the outcome of interest.

**Table 2.** Summary of the risk of bias assessment results.

| Authors | Was the Study's target population a close representation of the national population | Was the sampling frame a true or close representation of the target population | Was some form of random selection used to select the sample, OR, was a census undertaken | Was the likelihood of non-response bias minimal? | Were data collected directly from the subjects? | Was an acceptable case definition used in the study? | Was the study instrument shown to have reliability and validity | Was the same mode of data collection used for all subjects? | Was the length of the shortest prevalence period for the parameter of interest appropriate | Were the Numerator and denominator for the parameter of interest appropriate | Overall risk of bias |
| --- | --- | --- | --- | --- | --- | --- | --- | --- | --- | --- | --- |
| Ahmed et al., 2017 | High | Low | High | High | High | High | High | Low | Low | Low | Moderate |
| Aldington et al., 2014 | High | Low | Low | High | Low | Low | High | Low | Low | Low | Low |
| Bekrater et al., 2015 | Low | Low | High | High | Low | Low | High | High | Low | Low | Moderate |
| Bin Ayaz et al., 2015 | High | High | High | High | High | Low | High | High | High | Low | High |
| Bosmans et al., 2007 | High | Low | Low | High | Low | High | High | Low | Low | Low | Moderate |
| Buchanan et al., 1986 | High | High | High | High | Low | High | High | Low | High | Low | High |
| Byrne et al., 2011<sup>a</sup> | High | High | Low | High | Low | High | High | High | High | Low | High |
| Byrne et al., 2011<sup>b</sup> | High | High | Low | High | Low | High | High | High | High | Low | High |
| Clark et al., 2013 | High | Low | High | High | High | High | High | High | High | Low | High |
| Datta et al., 2004 | High | Low | High | Low | High | High | High | Low | High | Low | Moderate |
| Desmond et al., 2010 | High | Low | High | High | High | Low | High | Low | Low | Low | Moderate |
| Dijkstra et al., 2002 | High | Low | High | High | High | Low | High | Low | High | Low | Moderate |
| Ehde et al., 2000 | High | Low | Low | High | High | Low | High | Low | High | Low | Moderate |
| Ephraim et al., 2005 | Low | Low | Low | High | High | Low | High | Low | High | Low | Moderate |
| Gallagher et al., 2001 | High | High | High | High | High | High | High | Low | High | Low | High |
| Hanley et al., 2006 | High | Low | Low | High | High | Low | High | High | Low | Low | Moderate |
| Hanley et al., 2009 | High | High | Low | High | High | High | High | Low | High | Low | Moderate |
| Houghton et al., 1994 | High | Low | High | High | High | High | High | Low | High | Low | High |
| Hnoosh et al., 2014 | Low | High | Low | High | Low | Low | High | Low | Low | Low | Low |
| Kern et al., 2012 | Low | Low | Low | High | High | High | High | Low | High | Low | Moderate |
| Ketz et al., 2008 | High | Low | High | High | Low | Low | High | Low | High | Low | Moderate |
| Kooijman et al., 2000<sup>a</sup> | High | Low | High | Low | High | High | High | Low | High | Low | Moderate |
| Kooijman et al., 2000<sup>b</sup> | High | Low | High | Low | High | High | High | Low | High | Low | Moderate |
| Larbig et al., 2019 | High | High | High | High | Low | High | High | Low | Low | Low | Moderate |
| Morgan et al., 2017 | Low | Low | High | Low | High | High | High | Low | Low | Low | Moderate |
| Noguchi et al., 2019 | High | High | High | High | Low | Low | High | Low | Low | Low | Moderate |
| Penna et al., 2019 | High | High | Low | High | High | High | High | Low | High | Low | High |
| Probstner et al., 2010 | High | High | High | High | Low | High | High | High | High | Low | High |
| Rafferty et al., 2015 | High | High | High | Low | Low | High | High | Low | Low | Low | Moderate |
| Rahimi et al., 2012 | Low | Low | High | Low | Low | High | High | Low | Low | Low | Low |
| Rayegani et al., 2010 | High | Low | High | Low | Low | Low | High | Low | Low | Low | Low |
| Rasmus et al., 2017 | High | High | High | High | Low | High | High | Low | High | Low | High |
| Reiber et al.,<br>2010<sup>a</sup> | High | low | High | High | High | High | High | High | High | Low | High |
| Reiber et al.,<br>2010<sup>b</sup> | High | low | High | High | High | High | High | High | High | Low | High |
| Resnik et al.,<br>2019 | Low | Low | High | High | Low | High | High | Low | High | Low | Moderate |
| Richardson<br>et al., 2007 | High | Low | High | Low | Low | High | High | Low | High | Low | Moderate |
| Richardson<br>et al., 2015 | High | Low | High | High | Low | High | High | Low | Low | Low | Moderate |
| Schley et al.,<br>2008 | High | Low | High | High | High | High | High | High | High | Low | High |
| Wartan et<br>al., 1997 | High | Low | Low | Low | High | High | High | Low | High | Low | Moderate |
| Yin et al.,<br>2017 | High | Low | Low | High | High | High | High | Low | High | Low | Moderate |

### Prevalence of phantom limb pain

The estimates of PLP prevalence in people with limb amputations ranged between 0% and 85.6% (Kooijman, Dijkstra et al. 2000, Clark, Bowling et al. 2013), with most studies (31 out of 40) reporting a prevalence between 50% and 85.6% (Byrne 2011, Clark, Bowling et al. 2013). A prevalence of 0% was reported by the only included study that investigated PLP in adults with congenital amputations (Kooijman, Dijkstra et al. 2000). The pooling of all studies using a random effects model yielded an estimated prevalence of 63% [95% CI: 58.23-67.05], but with high statistical heterogeneity [I^2^=95.70% (95% CI: 95.10-96.20)] (Fig 3). The Egger’s regression analysis of all the included studies revealed no publication bias [-0.85 (95%CI: −4.13-2.43); p=0.60].

**Fig 3.**
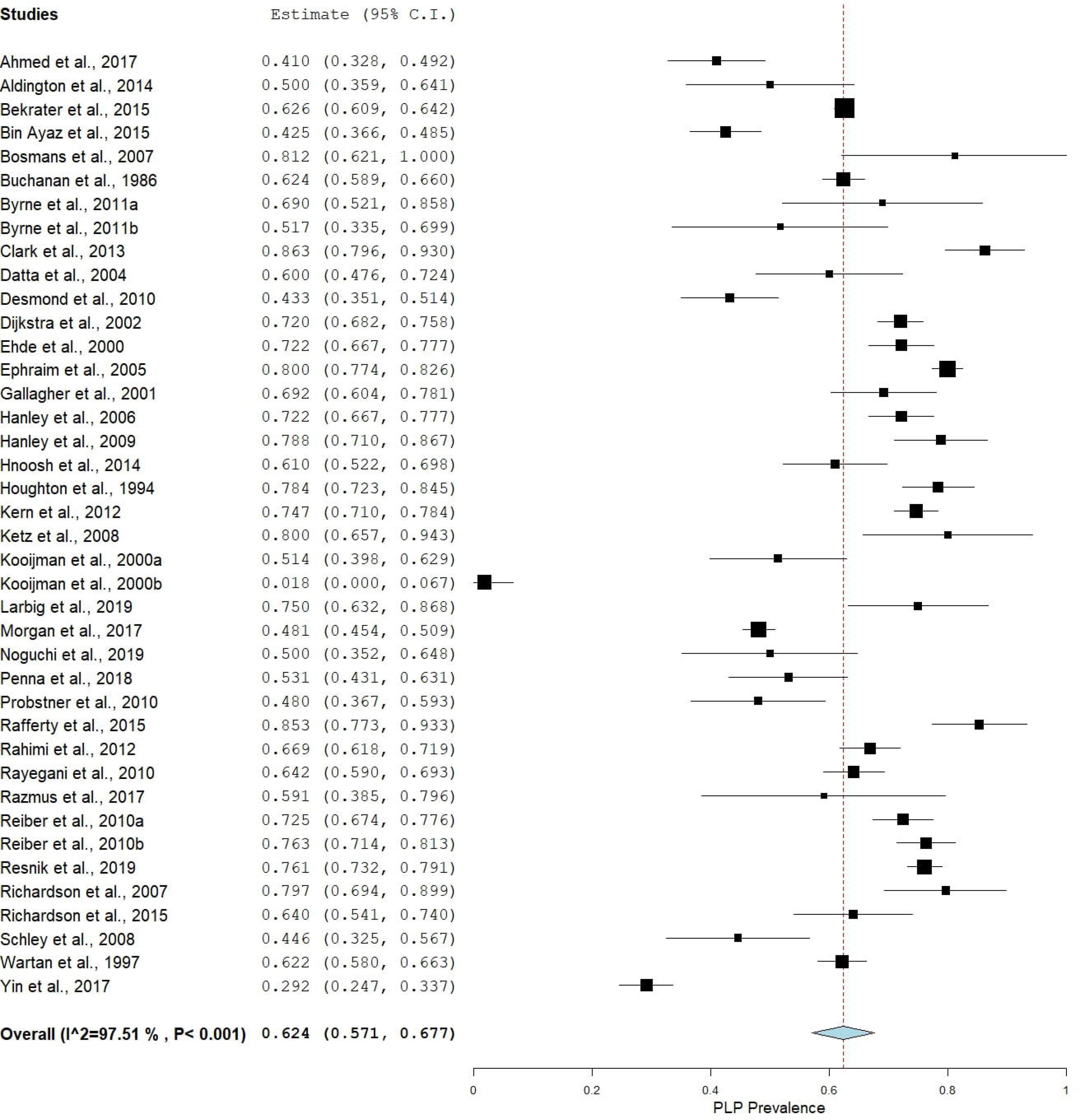
A forest plot showing the overall pooled estimated prevalence of PLP in people with amputations.

### Exploratory subgrouping

We were concerned by the high statistical heterogeneity in the primary meta-analysis, so we opted to deviate from protocol to conduct two exploratory meta-analyses with studies sub-grouped according to risk of bias score. The first exploratory subgroup analysis, including only the studies that scored low risk of bias overall, estimated prevalence at 63% [95% CI: 58.31-67.90] with moderate statistical heterogeneity [I^2^ =44.91 (95% CI: 43.90-45.20)] (Fig 4). The second exploratory subgroup analysis, including only the studies with moderate-high risk of bias, estimated prevalence at 63% [95% CI: 56.83-68.40], but with high statistical heterogeneity [I^2^ =97.75% (95% CI: 96.17-98.76)] (Fig 5). The Mann-Whitney U test that served as the sensitivity analysis for the effect of moderate-high risk of bias showed no difference between the estimated prevalence from these two meta-analyses [U=58.5, p=0.22].

**Fig 4.**
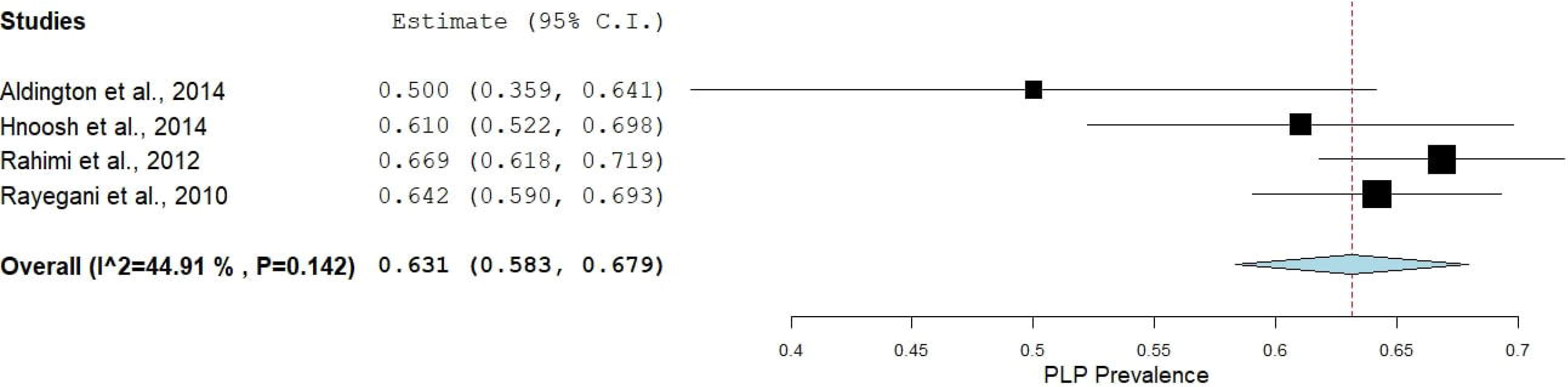
A subgroup analysis showing the pooled estimated prevalence of PLP in studies with low risk of bias.

**Fig 5.**
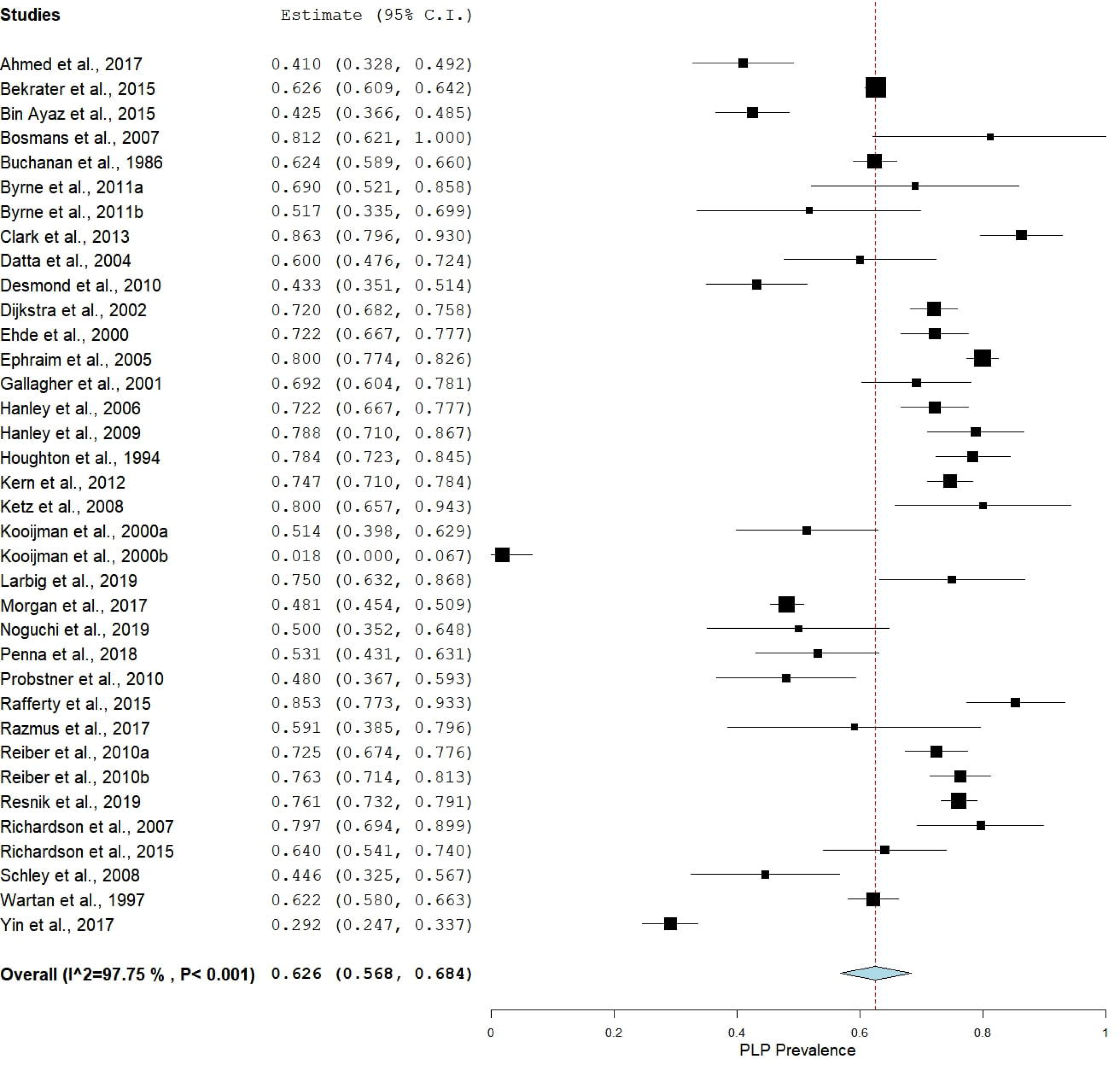
A subgroup analysis showing the pooled estimated prevalence of PLP in studies with moderate to high risk of bias.

The subgroup analyses stratified by the developmental status of the countries in which the studies were conducted showed an estimated pooled prevalence of 64.55% [95% CI: 59.62-69.34] in developed countries and 53.98% [95% CI: 44.79-63.05] in developing countries (Figs 6 & 7). The Mann-Whitney U test showed a statistically significant difference between the prevalence estimates of these two meta-analyses [U=57, p=0.04].

**Fig 6.**
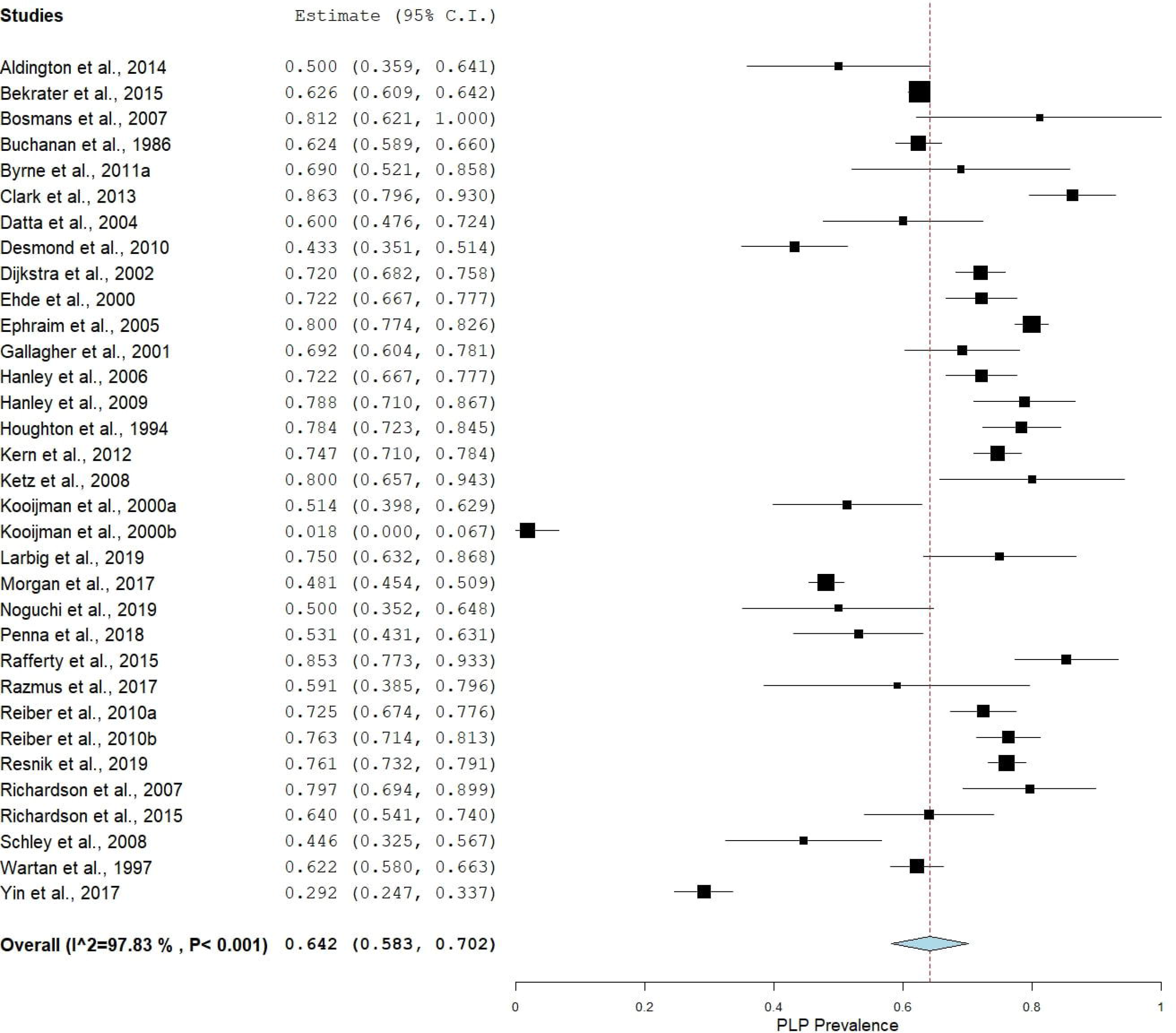
A subgroup analysis showing the pooled estimated prevalence of PLP in developed countries.

**Fig 7.**
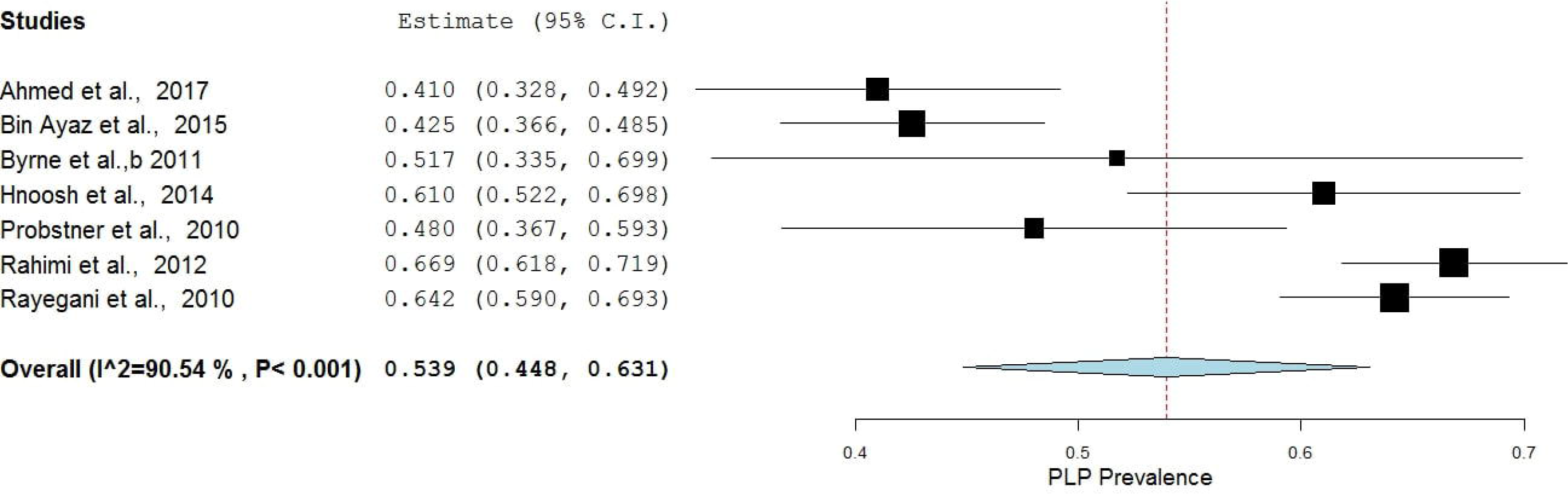
A subgroup analysis showing the pooled estimated prevalence of PLP in developing countries.

### Risk factors for phantom limb pain

Twenty-five potential risk factors had been studied in 15 studies representing 4102 participants. Of these, 10 pre-amputation, three peri-operative and eight post-amputation risk factors had data to support their positive association with PLP, and six pre-amputation, four peri-operative and three post-amputation risk factors had not been found to be positively associated with PLP. The risk factors found to be positively associated with PLP and their measures of association are summarised in Table 3.

**Table 3.** The summary of the risk factors for PLP and their measures of association with PLP.

| Author | Population | Outcome | Risk factor(s) | Measures of association | Strength of association |
| --- | --- | --- | --- | --- | --- |
| Ahmed et al., 2017 | Cancer patients who had undergone limb amputations | PLP | Post-amputation depression<br>Pre-amputation pain<br>Stump pain<br>Use of prosthesis<br>Sleep disturbance | 3.86 (1.75-8.53)<sup>‡</sup><br>2.83 (1.38-5.76)<sup>‡</sup><br>31.2 (8.97-108.50)<sup>‡</sup><br>2.83 (1.19-4.76)<sup>‡</sup><br>21.43 (8.28-55.43)<sup>‡</sup> | Strong<br>Moderate<br>Very strong<br>Moderate<br>Very strong |
| Buchanan et al., 1986 | Amputees who were receiving routine prosthetic services | PLP | Age | 0.12 (p<0.01)<sup>*</sup> | Weak |
| Desmond et al., 2010 | Members of the British Limbless Ex-Service Men's Association | PLP | Stump pain | 11.17 (p<0.01)<sup>‡</sup> | Very strong |
| Dijkstra et al., 2002 | Amputees who were receiving routine prosthetic services | PLP | Diabetic cause of amputation<br>Proximal site of amputation<br>Lower limb amputation<br>Bilateral amputations<br>Stump pain<br>Phantom sensations | 4 (p<0.001)<sup>‡</sup><br>1.60 (0.038)<sup>‡</sup><br>5.60 (p<0.001)<sup>‡</sup><br>8.20 (p=0.01)<sup>‡</sup><br>3.90 (p<0.001)<sup>‡</sup><br>19.50 (p<0.001)<sup>‡</sup> | Strong<br>Moderate<br>Strong<br>Strong<br>Strong<br>Very strong |
| Ephraim et al., 2005 | Amputees who had contacted the | PLP | Post-amputation depression | 2 (1.3-3.1)<sup>‡</sup><br>2.50 (1.3-4.7)<sup>‡</sup> | Moderate |
|  | Amputee Coalition of America (ACA) between 1998 and 2000 |  | Lower limb amputation<br>2 or more comorbidities<br>Widow | 2.70 (1.3-5.8)<sup>‡</sup><br>2.70 (1.1-6.5)<sup>‡</sup> | Moderate<br>Moderate<br>Moderate |
| Gallagher et al., 2001 | Amputees who were attending the Limb Fitting Clinic. | PLP | Proximal site of amputation<br>Traumatic cause of amputation<br>Sex (male)<br>Other medical problems<br>Lack of pre-amputation counselling | 15.65 (p<0.001)<sup>‡</sup><br>14.60 (p<0.002)<sup>‡</sup><br>3.76 (p<0.05)<sup>‡</sup><br>5.93 (p<0.02)<sup>‡</sup><br>4.74 (p<0.03)<sup>‡</sup> | Very strong<br>Very strong<br>Strong<br>Strong<br>Strong |
| Hanley et al., 2009 | Patients who had undergone upper-limb amputation 6 months or more before recruitment | PLP | Use of prosthesis | 4.23 (p<0.05)<sup>¶</sup> | Moderate |
| Hanley et al., 2006 | Patients who had undergone lower limb amputation | PLP | Pre-amputation pain<br>Stump pain | 0.48 (p<0.01)<sup>§</sup><br>0.53 (p<0.0001)<sup>§</sup> | Weak<br>Weak |
| Kooijman et al., 2000 <sup>a</sup> | Amputees using upper limb prosthesis | PLP | Phantom sensations<br>Stump pain | 11.30 (p=0.001)<sup>†</sup><br>1.90 (p=0.015)<sup>†</sup> | Very strong<br>Weak |
| Larbig et al., 2019 | Patients who had undergone upper or lower limb amputations | PLP | Pre-amputation depression<br>Pre-amputation pain<br>Stump pain | 2.05 (p<0.05)<sup>§</sup><br>4.22 (p<0.01)<sup>§</sup><br>3.90 (p<0.01)<sup>§</sup> | Moderate<br>Moderate<br>Moderate |
| Noguchi et al., 2019 | Patients who had undergone upper or lower limb amputations | PLP | Diabetic cause of amputation<br>Pre-amputation pain | 2.24 (p=0.032)<sup>‡</sup><br>6.36 (p=0.024)<sup>‡</sup> | Moderate<br>Strong |
| Razmus et al., 2017 | Occupants of the nursing home, and clients of the Public Institute of Orthopaedic Equipment | PLP | Phantom sensations | 4.94 (p<0.05)<sup>§</sup> | Strong |
| Richardson et al., 2007 | Patients who had undergone amputation of the lower limb due to peripheral vascular disease. | PLP | Stump pain<br>Increased ability to move the phantom limb.<br>Praying/hoping<br>Catastrophizing<br>Passive coping | 7.03 (1.34-36.82)<sup>‡</sup><br>8.31 (1.54-44.79)<sup>‡</sup><br>2.86 (1.68-13.18)<sup>‡</sup><br>3.28 (1.71-14.91)<sup>‡</sup><br>4.60 (6.50-25.00)<sup>‡</sup> | Strong<br>Strong<br>Moderate<br>Strong<br>Strong |
| Wartan et al., 1997 | Traumatic amputees | PLP | Phantom sensations | 107.30 (p<0.0001)<sup>§</sup> | Strong |
| Yin et al., 2017 | Amputees who underwent limb amputations at a tertiary hospital | PLP | Pre-amputation pain<br>Post-amputation epidural analgesia | 10.40 (p=0.002)<sup>‡</sup><br>4.90 (p=0.008)<sup>‡</sup> | Very strong<br>Strong |

¥ Point-biserial correlation analysis; ¶ Pearson's univariate correlation test; § Chi-squared; † Relative risk; ‡ Odds ratio

Lower limb amputation was positively associated with PLP (moderate to strong association) in two studies representing a total of 1450 participants (Dijkstra, Geertzen et al. 2002, Ephraim, Wegener et al. 2005). Stump pain was consistently positively associated with PLP (weak to very strong association) in seven studies representing a total of 1254 participants (Kooijman, Dijkstra et al. 2000, Dijkstra, Geertzen et al. 2002, Richardson, Glenn et al. 2006, Hanley, Jensen et al. 2007, Desmond and MacLachlan 2010, Ahmed, Bhatnagar et al. 2017, Larbig, Andoh et al. 2019). Phantom sensations were consistently positively associated with PLP (strong to very strong association) in four studies representing a total of 1156 participants (Wartan, Hamann et al. 1997, Kooijman, Dijkstra et al. 2000, Dijkstra, Geertzen et al. 2002, Razmus, Daniluk et al. 2017). Proximal site of amputation was positively associated with PLP (very strong association) in two studies representing a total of 604 participants (Gallagher, Butterworth et al. 1998, Dijkstra, Geertzen et al. 2002). Diabetic cause of amputation was positively associated with PLP (moderate to strong association) in two studies representing a total of 580 participants (Dijkstra, Geertzen et al. 2002, Noguchi, Saito et al. 2019). Persistent pre-amputation pain was positively associated with PLP in five studies representing a total of 881 participants (weak to very strong association) (Larbig, Montoya et al. 1996, Hanley, Jensen et al. 2007, Ahmed, Bhatnagar et al. 2017, Yin, Zhang et al. 2017, Noguchi, Saito et al. 2019) but was not associated with PLP in two studies representing a total of 625 participants. The risk factors which were not found to be positively associated with PLP are summarised in Table 4. Sex, age and traumatic cause of amputation were the most commonly assessed of these proposed risk factors. Sex was consistently not associated with PLP in six studies representing a total of 1836 participants (Kooijman, Dijkstra et al. 2000, Ephraim, Wegener et al. 2005, Dijkstra 2006, Hanley, Ehde et al. 2009, Ahmed, Bhatnagar et al. 2017, Noguchi, Saito et al. 2019). Age was not associated with PLP in three studies representing a total of 1062 participants (Ephraim, Wegener et al. 2005, Hanley, Ehde et al. 2009, Noguchi, Saito et al. 2019) but higher age was positively associated with PLP (weak association) in one study representing a total of 716 participants (Buchanan and Mandel 1986). A traumatic cause of amputation was not associated with PLP in two studies representing a total of 958 participants (Ephraim, Wegener et al. 2005, Noguchi, Saito et al. 2019) but was positively associated with PLP (very strong association) in one study representing a total of 104 participants (Gallagher, Allen et al. 2001). The meta-analysis of risk factors for PLP could not be conducted because of variations in methods of data collection and analysis across the included studies.

**Table 4.**
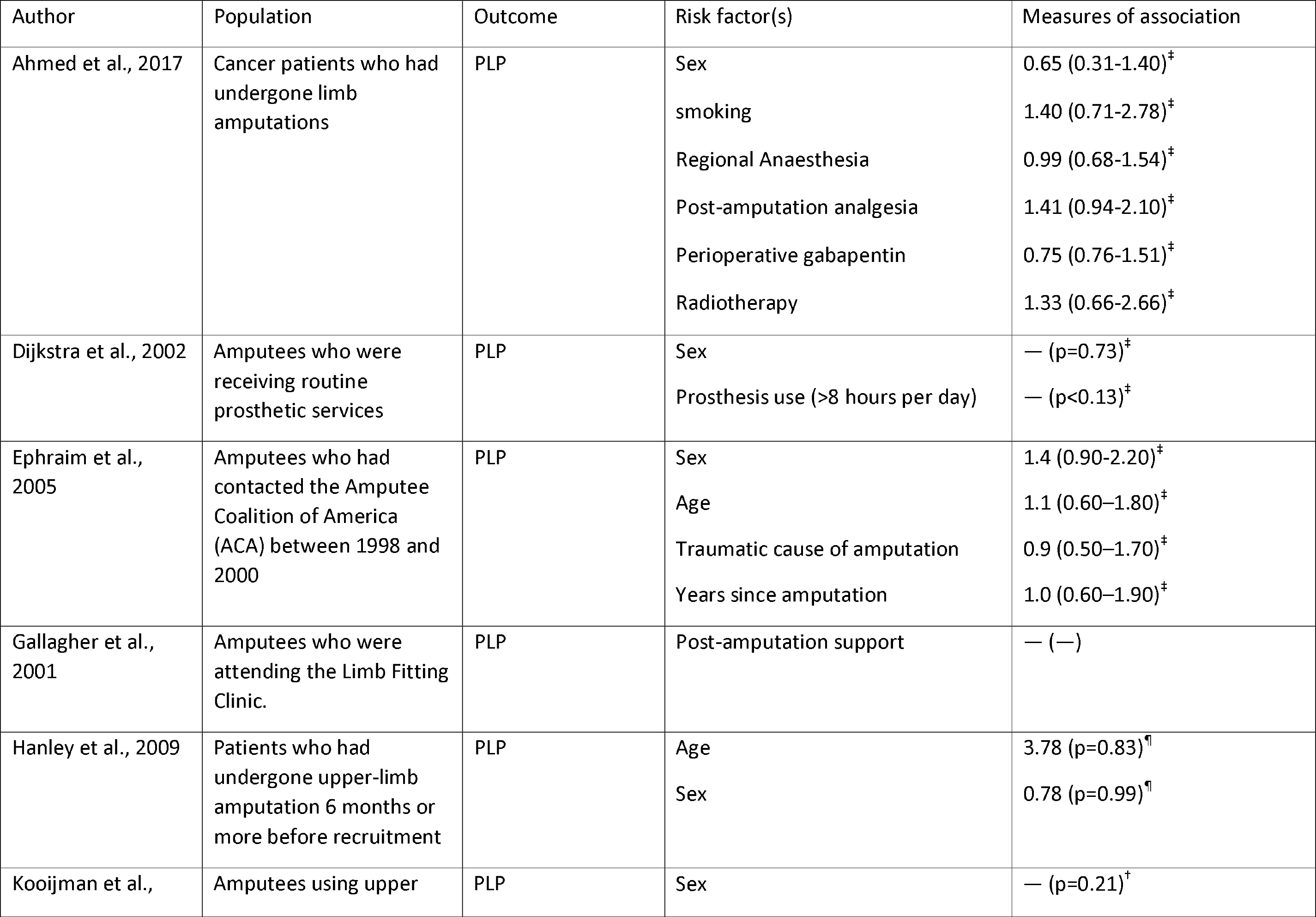
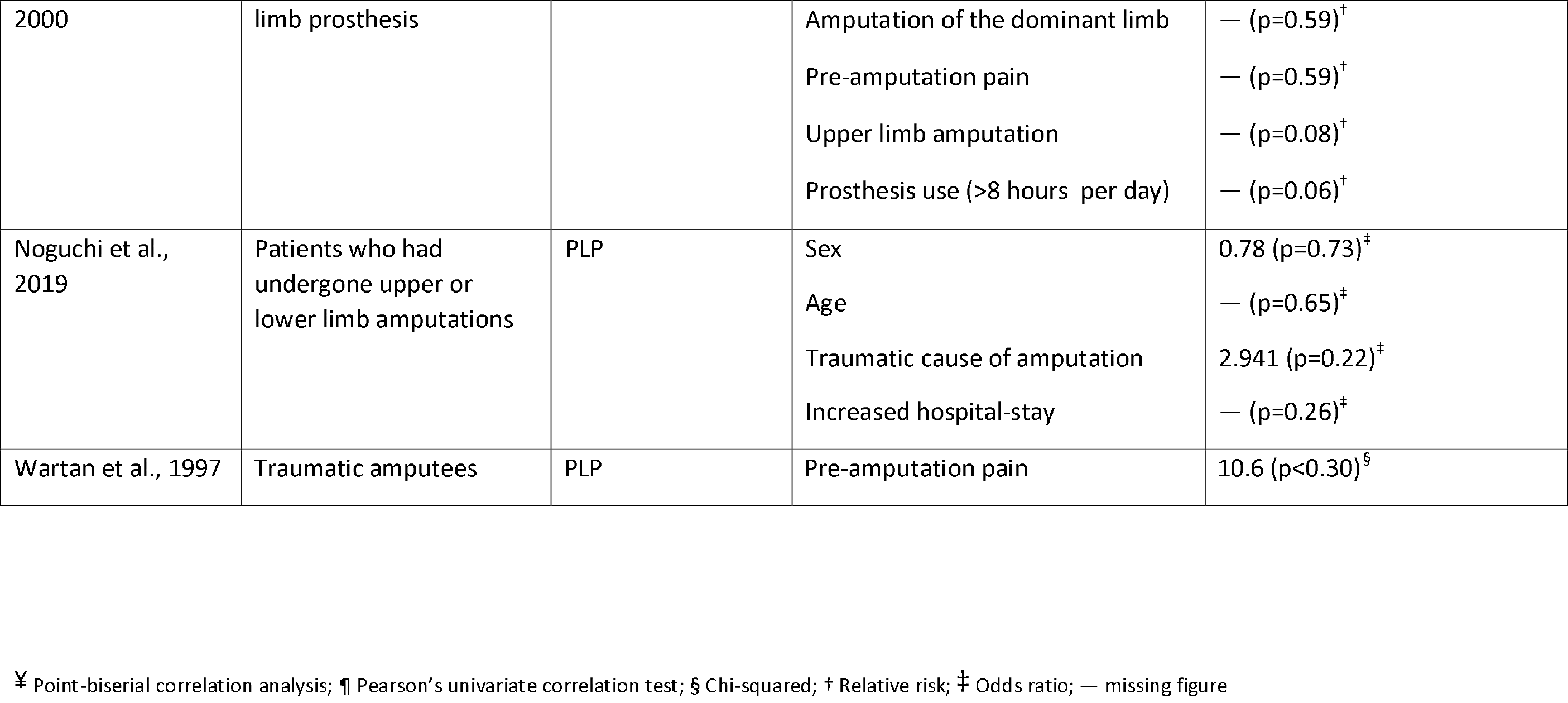
The summary of factors not associated with increased risk for PLP and their measures of association with PLP.

## Discussion

According to our knowledge, this is the first systematic review to pool the literature on the prevalence and risk factors for PLP in people with limb amputations. The results of this study estimate that PLP affects 63% of people with amputations. In addition, this study identified that lower limb amputation, stump pain, phantom sensations, persistent pre-amputation pain, proximal site of amputation and diabetic cause of amputation are risk factors for PLP.

### Phantom limb pain prevalence

The current primary meta-analysis estimated that 63% of people with amputations report PLP. This estimate suggests that approximately 8042 of 12765 participants in this study reported PLP. Interestingly, dividing studies by risk of bias revealed no difference in estimated prevalence, despite the ‘low risk of bias’ subgroup’s meta-analysis having lower statistical heterogeneity. In addition, the results of the Egger’s regression test indicated that the asymmetry of the funnel plot (Fig 8) was not significant (p=0.60), thus failing to suggest the presence of publication bias. Altogether, these findings suggest that the included studies provide a reasonably stable estimate of the prevalence of PLP in the population of people with amputations. The prevalence of PLP appears to be high, supporting that health professionals should be aware of the risk of this complication and that pragmatic interventions for preventing or alleviating PLP are needed.

**Fig 8.**
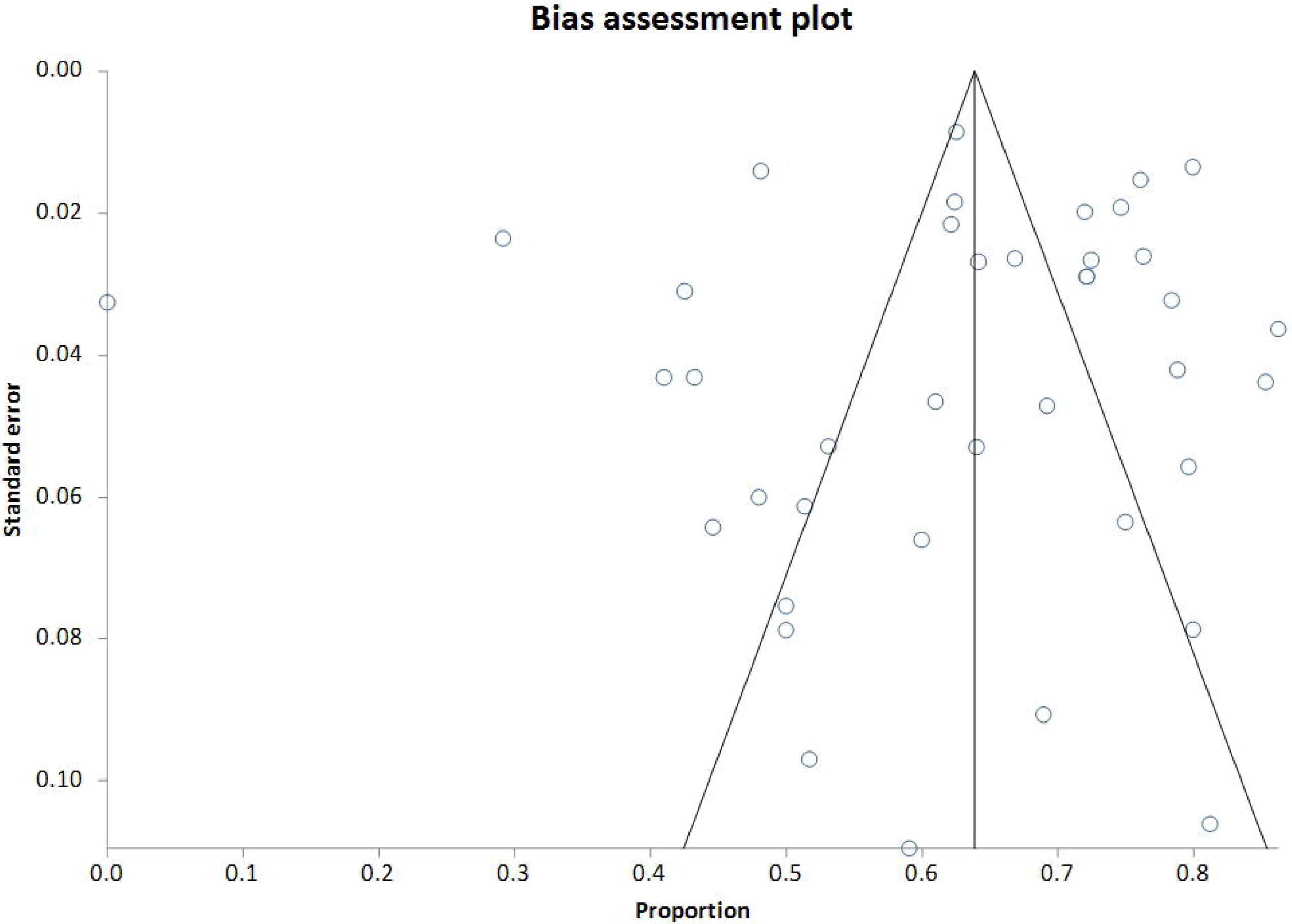
Funnel plot using data from the 40 included studies.

The meta-analysis that stratified the studies by country developmental status suggested that the prevalence of PLP was significantly lower in developing countries compared to developed countries [53.98% vs 64.55%; p=0.04]. This discrepancy is surprising and might be an artefact of selection bias linked either to the lower response rate seen in most of the studies conducted in developing countries, or to the strategy of recruiting participants from amongst patients receiving follow-up medical care. This recruitment strategy common in the developing country studies may have contributed to underestimation of PLP prevalence if amputees with PLP (in developing countries) without continuing medical care were excluded from samples, or may have contributed to an overestimation of PLP prevalence if having PLP made amputees more likely to remain in medical care. This uncertainty highlights the need to develop and implement novel recruitment strategies specific to people with amputations in developing countries so that they can be accounted for in future studies.

The included studies had varying risk of bias. However, the lack of statistically significant difference between the prevalence estimates from pooling of the studies with low risk of bias and pooling of the studies with moderate-high risk of bias suggests that the overall risk of bias in included studies had little impact on the prevalence of PLP. Nonetheless, the high risk of bias attributed to most studies for using an ambiguous definition of PLP (e.g pain felt in the limb after amputation) leaves the possibility that participants might have confused residual limb pain and PLP. We suspect that this might have resulted in an overestimation of the prevalence of PLP.

It is noteworthy that a prevalence of 0% was reported by the only study in this review that investigated PLP in adults with congenital amputations (Kooijman, Dijkstra et al. 2000). These findings are similar to those of other studies (not eligible for this review) that reported a low PLP prevalence of 7% (out of 57 patients) and 5.7% (out of 88 patients) in children with congenital amputations (Boonstra, Rijnders et al. 2000, Garcia, Flores et al. 2018). Although a robust conclusion cannot be drawn from three small studies, these findings suggest that people with congenital amputations may be less likely to experience PLP than those whose amputations were due to trauma or surgery (Gallagher, Allen et al. 2001). Perhaps the peripheral nerves severed during amputation play an important role in the initiation of PLP after amputation (Vaso, Adahan et al. 2014). In addition, the absence of pre-operative and peri-operative risk factors for PLP in this group might contribute to the low prevalence.

### Phantom limb pain risk factors

Five studies showed that PLP was more likely to occur in people who reported a history of persistent pre-operative pain than in those who did not report having had persistent limb pain prior to their amputation. One physiological mechanism that has been proposed to explain the link between pre-amputation pain and PLP is central sensitisation - where persistent pre-operative pain contributes to the hyperexcitability of the nervous system and functional changes in the cortical areas involved in the generation of pain (Lee, Zambreanu et al. 2008). These changes may continue to upregulate peripheral input after limb amputation, thus promoting PLP that shares the characteristics with pre-amputation pain (Rathmell and Kehlet 2011). In fact, over 60% of the patients who experienced persistent pre-amputation pain reported similar characteristics of their PLP (Jensen, Krebs et al. 1985, Katz and Melzack 1990). This apparent relationship highlights the importance of addressing limb pain very early in patients who are at high risk of having their limbs amputated. The early and effective management of pre-amputation pain may reduce risk of developing PLP and improve physical and psychological outcomes often related to delayed or ineffective management of PLP (Hanley, Jensen et al. 2004, Hanley, Jensen et al. 2007)

Two studies showed that PLP was more likely to occur after lower limb amputation than after upper limb amputation (Ephraim, Wegener et al. 2005, Dijkstra 2006). The authors proposed that the use of a cosmetic prosthetic leg, rather than a prosthesis that provided sensory input was a likely contributor to pain in people with lower limb amputations since 70%-78.8% of cosmetic prosthetic leg users had PLP. Lack of proprioceptive feedback during the use of a prosthetic leg has been linked to poor motor control, possibly leading to stump irritation that may trigger PLP (Morgan, Friedly et al. 2017, Page, George et al. 2018). This proposed link is partially supported by seven studies in this review which suggested that PLP was more likely to occur in people with stump pain than in those without stump pain (Kooijman, Dijkstra et al. 2000, Dijkstra, Geertzen et al. 2002, Hanley, Ehde et al. 2006, Richardson, Glenn et al. 2006, Desmond and MacLachlan 2010, Ahmed, Bhatnagar et al. 2017, Larbig, Andoh et al. 2019). Interestingly, Dietrich and colleagues investigated the effects of a leg prosthesis with somatosensory feedback on pain and lower limb function (Dietrich, Nehrdich et al. 2018). In that study, participants used prosthetic legs with pressure sensors that provided comfortable electrical feedback to the patient’s thigh whenever the prosthetic foot touched the ground. At the end of two weeks of training, the participants had improved function of the lower limb and reduced severity and frequency of PLP. Further, the patients reported greater satisfaction, longer walking distances and improved dynamic stability than prior to the training. These results suggest that people with lower limb amputations might benefit more from using a prosthetic leg with somatosensory feedback than from using a cosmetic prosthesis. However, the mechanisms by which prosthetic legs with somatosensory feedback reduce PLP are not clear. Therefore, it would be interesting to investigate the mechanisms by which somatosensory feedback from a prosthetic leg might influence PLP.

Four studies showed that PLP was more likely to occur in amputees with non-painful phantom sensations than in those without non-painful phantom sensations (Wartan, Hamann et al. 1997, Kooijman, Dijkstra et al. 2000, Razmus, Daniluk et al. 2017). In these studies, 70%-100% of amputees with phantom sensations also had PLP. The co-occurrence of these post-amputation sensations suggest that they may share neural mechanisms with PLP (Razmus, Daniluk et al. 2017). An fMRI study by Andoh et al showed that inducing non-painful phantom sensations in people with amputations activated the somatosensory and premotor cortices contralateral to the amputated limb (Andoh, Diers et al. 2017). The activation of similar cortical areas has been recorded in patients with PLP upon induction of their PLP (Flor and Elbert 1995, Lotze, Montoya et al. 1999, Karl, Birbaumer et al. 2001, Lotze, Flor et al. 2001, Flor, Nikolajsen et al. 2006). The similarities in cortical activation patterns might explain a link between PLP and non-painful phantom sensations.

Two studies showed that PLP was more likely to occur in people with proximal amputations than in those with distal amputations (Gallagher, Allen et al. 2001, Dijkstra, Geertzen et al. 2002). These findings line up with a narrative review that reported an increase in the incidence of PLP with more proximal amputations (Manchikanti and Singh 2004). Proximal amputations are associated with an increased risk of failure of wound healing, which may result in infection or stump pain (Stone, Flaherty et al. 2006). However, the reasons why proximal amputations should be more likely to lead to PLP than distal amputations are not clear (Kelle, Kozanoglu et al. 2017).

Another interesting finding was that the lack of pre-amputation counselling was positively associated with PLP (strong association) in a study representing a total of 104 participants (Gallagher, Allen et al. 2001). This suggests that patients who receive counselling prior to their amputation maybe less likely to report PLP compared to those who do not receive counselling. The exact details of the counselling were not reported. However, the strong association between the pre-operative counselling and pain reduction highlights the importance of pre-amputation counselling on PLP. No other study has specifically identified pre-operative counselling as a predictor of decreased PLP after limb amputation. However, the results of a narrative review suggest that pre-operative counselling may improve outcomes in patients undergoing various forms of surgery (Powell, Scott et al. 2016).

### Limitations

The sample in this systematic review was skewed towards males, in that 9826 (81.73%) of the 12022 participants were male. Therefore, the results might not hold for females. We could not perform a subgroup analysis by sex because we did not have individual patient data, nor was analysis by sex an objective identified in the protocol. However, the data on risk factors provide no support for sex influencing the likelihood of PLP after amputation. It was not possible to conduct a meta-analysis on the risk factors for PLP because the included studies used varying methodological approaches and measures of association. None of the included studies used an outcome measure that has been validated for assessing PLP. In fact, we are not aware of any instrument that has been validated for assessing PLP. Such a standardised tool for assessing PLP would be useful to provide us with reliable data. Most studies in this review had moderate-high risk of bias. There is a clear need for high-quality studies to raise the credibility of future meta-analyses. Finally, the search strategy for this study was designed specifically to identify prevalence studies. Therefore, although we did conduct an exploratory search for additional studies of risk factors for PLP, there is a possibility that we could have missed some studies that investigated risk factors for PLP if they did not also estimate PLP prevalence. In consideration of this possibility, the review of risk factors was classified as an exploratory analysis. It is important to note that the results of this systematic review were derived from studies conducted mostly in Europe, North-America and Asia. To the best of our knowledge, no study has been conducted in Africa, and research in this area is indicated to inform us about the burden and risk factors for PLP in the African population.

### Conclusion

This systematic review and meta-analysis estimates that six of every 10 people with an amputation report PLP – a high and important prevalence of PLP. Health care professionals ought to be aware of the high rates of PLP and implement strategies to reduce PLP by addressing known risk factors, specifically those identified by the current study. Stump pain and post-amputation depression are all known and modifiable risk factors that are consistently positively associated with PLP. Awareness of these risk factors may motivate health care professionals to address them early in treatment to prevent the onset of PLP in people with amputations.

## Data Availability

The data will be made available to anyone upon reasonable request.

## Acknowledgements

The authors thank Mrs Mary Shelton (Health Sciences reference librarian, University of Cape Town) for assisting with the development of the search strategy.

## Notes

### Competing Interest Statement

The authors have declared no competing interest.

### Funding Statement

The first author is supported by the start-up development grant of the University of Cape Town

### Author Declarations

All relevant ethical guidelines have been followed and any necessary IRB and/or ethics committee approvals have been obtained.

Any clinical trials involved have been registered with an ICMJE-approved registry such as ClinicalTrials.gov and the trial ID is included in the manuscript.

## References

Ahmed, A., S. Bhatnagar, S. Mishra, D. Khurana, S. Joshi and S. M. Ahmad (2017). “Prevalence of phantom limb pain, stump pain, and phantom limb sensation among the amputated cancer patients in India: A prospective, observational study.” Indian journal of palliative care 23(1): 24.

Aldington, D., C. Small, D. Edwards, J. Ralph, P. Woods, S. Jagdish and R. A. Moore (2014). “A survey of post-amputation pains in serving military personnel.” J R Army Med Corps 160(1): 38–41.

Allen, M. (2017). The SAGE encyclopedia of communication research methods, SAGE Publications.

Andoh, J., M. Diers, C. Milde, C. Frobel, D. Kleinböhl and H. Flor (2017). “Neural correlates of evoked phantom limb sensations.” Biological Psychology 126: 89–97.

Bekrater-Bodmann, R., M. Schredl, M. Diers, I. Reinhard, J. Foell, J. Trojan, X. Fuchs and H. Flor (2015). “Post-amputation pain is associated with the recall of an impaired body representation in dreams-results from a nation-wide survey on limb amputees.” PLoS One 10(3): e0119552.

Bin Ayaz, S., M. Ikram, S. Matee, A. A. Khan, M. Ahmad and M. Fahim (2015). “FREQUENCY AND THE RELATED SOCIO-DEMOGRAPHIC AND CLINICAL FACTORS OF PHANTOM LIMB PAIN IN TRAUMATIC AMPUTEES PRESENTING AT A TERTIARY CARE REHABILITATION SETUP.” Pakistan Armed Forces Medical Journal(6): 782–788.

Boonstra, A. M., L. J. Rijnders, J. W. Groothoff and W. H. Eisma (2000). “Children with congenital deficiencies or acquired amputations of the lower limbs: functional aspects.” Prosthet Orthot Int 24(1): 19–27.

Buchanan, D. C. and A. R. Mandel (1986). “The prevalence of phantom limb experience in amputees.” Rehabilitation Psychology 31(3): 183.

Byrne, K. P. (2011). “Survey of phantom limb pain, phantom sensation and stump pain in Cambodian and New Zealand amputees.” Pain Med 12(5): 794–798.

Clark, R. L., F. L. Bowling, F. Jepson and S. Rajbhandari (2013). “Phantom limb pain after amputation in diabetic patients does not differ from that after amputation in nondiabetic patients.” Pain 154(5): 729–732.

Cohen, J. (1960). “A coefficient of agreement for nominal scales.” Educational and psychological measurement 20(1): 37–46.

Datta, D., K. Selvarajah and N. Davey (2004). “Functional outcome of patients with proximal upper limb deficiency -- acquired and congenital.” Clinical Rehabilitation 18(2): 172–177.

Desmond, D. M. and M. MacLachlan (2010). “Prevalence and characteristics of phantom limb pain and residual limb pain in the long term after upper limb amputation.” International Journal of Rehabilitation Research 33(3): 279–282.

Dietrich, C., S. Nehrdich, S. Seifert, K. R. Blume, W. H. R. Miltner, G. O. Hofmann and T. Weiss (2018). “Leg Prosthesis With Somatosensory Feedback Reduces Phantom Limb Pain and Increases Functionality.” Frontiers in Neurology 9(270).

Dijkstra, P. U. (2006). “’Re: Phantom limb pain’: Commentary reply.” Journal of Pain and Symptom Management 32(2): 103–103.

Dijkstra, P. U., H. B. Geertzen, R. Stewart and C. P. van der Schans (2002). “Phantom Pain and Risk Factors: A Multivariate Analysis.” Journal of Pain and Symptom Management 24(6): 578–585.

Egger, M., G. D. Smith, M. Schneider and C. Minder (1997). “Bias in meta-analysis detected by a simple, graphical test.” Bmj 315(7109): 629-634.

Ehde, D. M., J. M. Czerniecki, D. G. Smith, K. M. Campbell, W. T. Edwards, M. P. Jensen and L. R. Robinson (2000). “Chronic phantom sensations, phantom pain, residual limb pain, and other regional pain after lower limb amputation.” Archives of physical medicine and rehabilitation 81(8): 1039–1044.

Ephraim, P. L., S. T. Wegener, E. J. MacKenzie, T. R. Dillingham and L. E. Pezzin (2005). “Phantom pain, residual limb pain, and back pain in amputees: results of a national survey.” Archives of Physical Medicine & Rehabilitation 86(10): 1910–1919.

Flor, H. and T. Elbert (1995). “Phantom-limb pain as a perceptual correlate of cortical reorganization following arm amputation.” Nature 375(6531): 482.

Flor, H., L. Nikolajsen and T. S. Jensen (2006). “Phantom limb pain: a case of maladaptive CNS plasticity?” Nature Reviews Neuroscience 7(11): 873.

Frankfort-Nachmias, C. and A. Leon-Guerrero (2017). Social statistics for a diverse society, Sage Publications.

Gagnier, J. J., D. Moher, H. Boon, J. Beyene and C. Bombardier (2012). “Investigating clinical heterogeneity in systematic reviews: a methodologic review of guidance in the literature.” BMC medical research methodology 12(1): 111.

Gallagher, P., D. Allen and M. MacLachlan (2001). “Phantom limb pain and residual limb pain following lower limb amputation: a descriptive analysis.” Disability & Rehabilitation 23(12): 522–530.

Gallagher, S., G. E. Butterworth, A. Lew and J. Cole (1998). “Hand-mouth coordination, congenital absence of limb, and evidence for innate body schemas.” Brain Cogn 38(1): 53–65.

Garcia, D., E. Flores, P. Nahuelhual and F. Solis (2018). “Phantom pain in congenital amputees: Myth or reality?” Annals of Physical and Rehabilitation Medicine 61: e118.

Gosselin, R. A., Y. A. Gyamfi and S. Contini (2011). “Challenges of meeting surgical needs in the developing world.” World J Surg 35(2): 258–261.

Hanley, M. A., D. M. Ehde, K. M. Campbell, B. Osborn and D. G. Smith (2006). “Self-reported treatments used for lower-limb phantom pain: descriptive findings.” Archives of Physical Medicine & Rehabilitation 87(2): 270–311.

Hanley, M. A., D. M. Ehde, M. Jensen, J. Czerniecki, D. C. Smith and L. R. Robinson (2009). “Chronic Pain Associated with Upper-Limb Loss.” American Journal of Physical Medicine & Rehabilitation 88(9): 742–754.

Hanley, M. A., M. P. Jensen, D. M. Ehde, A. J. Hoffman, D. R. Patterson and L. R. Robinson (2004). “Psychosocial predictors of long-term adjustment to lower-limb amputation and phantom limb pain.” Disability & Rehabilitation 26(14/15): 882-893.

Hanley, M. A., M. P. Jensen, D. G. Smith, D. M. Ehde, W. T. Edwards and L. R. Robinson (2007). “Preamputation Pain and Acute Pain Predict Chronic Pain After Lower Extremity Amputation.” The Journal of Pain 8(2): 102–109.

Healey, J. F. (2014). Statistics: A tool for social research, Cengage Learning.

Higgins, J. P., S. G. Thompson, J. J. Deeks and D. G. Altman (2003). “Measuring inconsistency in meta-analyses.” BMJ: British Medical Journal 327(7414): 557.

Hnoosh, A. H. (2014). “Phantom Limb and pain after traumatic lower extremity amputation.” Journal of the Faculty of Medicine 56(1): 57–61.

Hoy, D., P. Brooks, A. Woolf, F. Blyth, L. March, C. Bain, P. Baker, E. Smith and R. Buchbinder (2012). “Assessing risk of bias in prevalence studies: modification of an existing tool and evidence of interrater agreement.” Journal of clinical epidemiology 65(9): 934–939.

Jensen, T. S., B. Krebs, J. Nielsen and P. Rasmussen (1985). “Immediate and long-term phantom limb pain in amputees: incidence, clinical characteristics and relationship to pre-amputation limb pain.” Pain 21(3): 267–278.

Karl, A., N. Birbaumer, W. Lutzenberger, L. G. Cohen and H. Flor (2001). “Reorganization of motor and somatosensory cortex in upper extremity amputees with phantom limb pain.” Journal of Neuroscience 21(10): 3609–3618.

Katz, J. and R. Melzack (1990). “Pain ‘memories’ in phantom limbs: Review and clinical observations.” Pain 43(3): 319–336.

Kelle, B., E. Kozanoglu, O. S. Bicer and I. Tan (2017). “Association between phantom limb complex and the level of amputation in lower limb amputee.” Acta Orthop Traumatol Turc 51(2): 142–145.

Kern, U., V. Busch, M. Rockland, M. Kohl and F. Birklein (2009). “[Prevalence and risk factors of phantom limb pain and phantom limb sensations in Germany. A nationwide field survey].” Schmerz 23(5): 479–488.

Ketz, A. K. (2008). “The experience of phantom limb pain in patients with combat-related traumatic amputations.” Archives of Physical Medicine & Rehabilitation 89(6): 1127–1132.

Kooijman, C. M., P. U. Dijkstra, J. H. Geertzen, A. Elzinga and C. P. van der Schans (2000). “Phantom pain and phantom sensations in upper limb amputees: an epidemiological study.” Pain 87(1): 33–41.

Larbig, W., J. Andoh, E. Huse, D. Stahl-Corino, P. Montoya, Z. e. Seltzer and H. Flor (2019). “Pre-and postoperative predictors of phantom limb pain.” Neuroscience letters 702: 44–50.

Larbig, W., P. Montoya, H. Flor, H. Bilow, S. Weller and N. Birbaumer (1996). “Evidence for a change in neural processing in phantom limb pain patients.” Pain 67(2-3): 275-283.

Lee, M. C., L. Zambreanu, D. K. Menon and I. Tracey (2008). “Identifying brain activity specifically related to the maintenance and perceptual consequence of central sensitization in humans.” J Neurosci 28(45): 11642–11649.

Limakatso, K., G. J. Bedwell, V. J. Madden and R. Parker (2019). “The prevalence of phantom limb pain and associated risk factors in people with amputations: a systematic review protocol.” Systematic reviews 8(1): 17–17.

Lotze, M., H. Flor, W. Grodd, W. Larbig and N. Birbaumer (2001). “Phantom movements and pain An fMRI study in upper limb amputees.” Brain 124(11): 2268–2277.

Lotze, M., P. Montoya, M. Erb, E. Hülsmann, H. Flor, U. Klose, N. Birbaumer and W. Grodd (1999). “Activation of cortical and cerebellar motor areas during executed and imagined hand movements: an fMRI study.” Journal of cognitive neuroscience 11(5): 491–501.

Maimela, E., M. Alberts, S. E. Modjadji, S. S. Choma, S. A. Dikotope, T. S. Ntuli and J.-P. Van Geertruyden (2016). “The prevalence and determinants of chronic non-communicable disease risk factors amongst adults in the Dikgale health demographic and surveillance system (HDSS) site, Limpopo Province of South Africa.” PLoS One 11(2): e0147926.

Manchikanti, L. and V. Singh (2004). “Managing phantom pain.” Pain Physician 7(3): 365–375.

Mishra, S., S. Bhatnagar, D. Gupta and A. Diwedi (2008). “Incidence and management of phantom limb pain according to World Health Organization analgesic ladder in amputees of malignant origin.” American Journal of Hospice and Palliative Medicine® 24(6): 455–462.

Moher, D., L. Shamseer, M. Clarke, D. Ghersi, A. Liberati, M. Petticrew, P. Shekelle, L. A. Stewart and P.-P. Group (2015). “Preferred reporting items for systematic review and meta-analysis protocols (PRISMA-P) 2015 statement.” Systematic Reviews 4(1): 1.

Moloi, A. H., D. Watkins, M. E. Engel, S. Mall and L. Zühlke (2016). “Epidemiology, health systems and stakeholders in rheumatic heart disease in Africa: a systematic review protocol.” BMJ open 6(5): e011266.

Morgan, S. J., J. L. Friedly, D. Amtmann, R. Salem and B. J. Hafner (2017). “Cross-Sectional Assessment of Factors Related to Pain Intensity and Pain Interference in Lower Limb Prosthesis Users.” Arch Phys Med Rehabil 98(1): 105–113.

Neil, M. (2015). “Pain after amputation.” BJA Education 16(3): 107–112.

Nicolich, M. J. and J. F. Gamble (2011). “What is the minimum risk that can be estimated from an epidemiology Study.” Advanced Topics in Environmental Health and Air Pollution Case Studies: 3-26.

Noguchi, S., J. Saito, K. Nakai, M. Kitayama and K. Hirota (2019). “Factors affecting phantom limb pain in patients undergoing amputation: retrospective study.” Journal of anesthesia 33(2): 216–220.

Page, D. M., J. A. George, D. T. Kluger, C. Duncan, S. Wendelken, T. Davis, D. T. Hutchinson and G. A. Clark (2018). “Motor Control and Sensory Feedback Enhance Prosthesis Embodiment and Reduce Phantom Pain After Long-Term Hand Amputation.” Frontiers in Human Neuroscience 12.

Powell, R., N. W. Scott, A. Manyande, J. Bruce, C. Vogele, L. M. Byrne-Davis, M. Unsworth, C. Osmer and M. Johnston (2016). “Psychological preparation and postoperative outcomes for adults undergoing surgery under general anaesthesia.” Cochrane Database Syst Rev(5): Cd008646.

Rafferty, M., T. M. Bennett Britton, B. T. Drew and R. D. Phillip (2015). “Cross-sectional study of alteration of phantom limb pain with visceral stimulation in military personnel with amputation.” J Rehabil Res Dev 52(4): 441–448.

Rahimi, A., B. Mousavi, M. Soroush, M. Masumi and A. Montazeri (2012). “Pain and health-related quality of life in war veterans with bilateral lower limb amputations.” Trauma Monthly 17(2): 282–286.

Rathmell, J. P., M.D. and H. Kehlet, M.D., Ph.D. (2011). “Do We Have the Tools to Prevent Phantom Limb Pain?” Anesthesiology: The Journal of the American Society of Anesthesiologists 114(5): 1021–1024.

Rathvon, D. (2017). “EndNote X8--Citation Manager--What’s New?".

Rayegani, S. M., A. Aryanmehr, M. R. Soroosh and M. Baghbani (2010). “Phantom pain, phantom sensation, and spine pain in bilateral lower limb amputees: Results of a national survey of Iraq-Iran war victims’ health status.” Journal of Prosthetics and Orthotics 22(3): 162–165.

Razmus, M., B. Daniluk and P. Markiewicz (2017). “Phantom limb phenomenon as an example of body image distortion.” Current Problems of Psychiatry 18(2): 153–159.

Reiber, G. E., L. V. McFarland, S. Hubbard, C. Maynard, D. K. Blough, J. M. Gambel and D. G. Smith (2010). “Servicemembers and veterans with major traumatic limb loss from Vietnam war and OIF/OEF conflicts: Survey methods, participants, and summary findings.” Journal of Rehabilitation Research & Development 47(3): 275–297.

Richardson, C., S. Glenn, T. Nurmikko and M. Horgan (2006). “Incidence of phantom phenomena including phantom limb pain 6 months after major lower limb amputation in patients with peripheral vascular disease.” The Clinical journal of pain 22(4): 353–358.

Rosenthal, J. A. (1996). “Qualitative descriptors of strength of association and effect size.” Journal of social service Research 21(4): 37–59.

Sherman, R. A., C. J. Sherman and N. G. Gall (1980). “A survey of current phantom limb pain treatment in the United States.” Pain 8(1): 85–99.

Stone, P. A., S. K. Flaherty, A. F. AbuRahma, S. M. Hass, J. M. Jackson, J. D. Hayes, M. J. Hofeldt, C. S. Hager and M. S. Elmore (2006). “Factors affecting perioperative mortality and wound-related complications following major lower extremity amputations.” Annals of vascular surgery 20(2): 209–216.

Vaso, A., H.-M. Adahan, A. Gjika, S. Zahaj, T. Zhurda, G. Vyshka and M. Devor (2014). “Peripheral nervous system origin of phantom limb pain.” PAIN® 155(7): 1384–1391.

Ventham, N., P. Heyburn and N. Huston (2010). “Prevalence of phantom limb pain in diabetic and non-diabetic leg amputees: a cross-sectional observational survey.” European Journal of Pain Supplements 4(1): 106–107.

Wartan, S. W., W. Hamann, J. R. Wedley and I. McColl (1997). “Phantom pain and sensation among British veteran amputees.” Br J Anaesth 78(6): 652–659.

Weeks, S. R., V. C. Anderson-Barnes and J. W. Tsao (2010). “Phantom limb pain: theories and therapies.” The neurologist 16(5): 277–286.

WESP. (2018). “Data sources, country classifications and aggregation methodology. (2014, January 25).” (2014, January 25), from Retrieved from http://www.un.org/en/development/desa/policy/wesp/wesp_current/2014wesp_country_classification.pdf.

Yin, Y., L. Zhang, H. Xiao, C.-B. Wen, Y.-E. Dai, G. Yang, Y.-X. Zuo and J. Liu (2017). “The pre-amputation pain and the postoperative deafferentation are the risk factors of phantom limb pain: a clinical survey in a sample of Chinese population.” BMC anesthesiology 17(1): 69.

